# Extending upon: What effect might border screening have on preventing importation of COVID-19 compared with other infections? – Considering the additional effect of post-arrival isolation

**DOI:** 10.1101/2021.12.23.21268332

**Authors:** Declan Bays, Emma Bennett, Thomas Finnie

## Abstract

We recently described a simple model through which we assessed what effect subjecting travellers to a single on-arrival test might have on reducing risk of importing disease cases during simulated outbreaks of COVID-19, Influenza, SARS, and Ebola. We build upon this work to allow for the additional requirement that inbound travellers also undergo a period of self-isolation upon arrival, where upon completion the traveller is again tested for signs of infection prior to admission across the border. Prior results indicated that a single on-arrival test has the potential to detect 9% of travellers infected with COVID-19, compared to 35%, 10% and 3% for travellers infected with influenza, SARS, and Ebola respectively. Our extended model shows that testing administered after a 2-day isolation period may be able to detect up to 41%, 97%, 44% and 15% of COVID-19, Influenza, SARS, and Ebola infected travellers respectively. Longer self-isolation periods increase detection rates further, with an 8-day self-isolation period suggesting detection rates of up to 94%, 100%, 98% and 62% for travellers infected with COVID-19, Influenza, SARS, and Ebola respectively. These results therefore suggest that testing arrivals after an enforced period of self-isolation may present a reasonable method of protecting against case importation during international outbreaks.

## Introduction

In a recent paper[1], we described an adaptable model which could be used to assess the effectiveness of implementing a strict policy of on-arrival testing at airports during international disease outbreaks. We subsequently used this model to evaluate the probability that a single test at-point-of-arrival would be able to detect infected travellers across a range of scenarios, given that they were not detected by assumed exit screening (i.e. having not fully incubated by the time they boarded their flight). Scenarios were defined by the disease that travellers were infected with, the exposure window (the time prior to departure within which travellers could have become infected), and whether travellers took a short, medium, or long-haul flight. Results showed that in the best-case scenario, screening has the capability to detect 8.8% of travellers infected with COVID-19 compared to 34.8.%, 9.7% and 3.0% for travellers infected with influenza, SARS, and Ebola respectively. This suggests that screening at point of arrival alone is unlikely to provide an adequate level of protection from international importations.

Intuition would reason that these detection rates are low since the time between pre-departure and post-arrival testing is not enough to allow a significant proportion of the infected travellers to incubate, and thus become detectable. We would therefore expect to raise these rates of detection by increasing the time between the pre-departure and post-arrival testing, allowing addition time for infections to complete their incubation period, and hence be picked up by subsequent testing. This can be achieved by requiring new arrivals to self-isolate for a fixed period prior to the administering of the post-arrival test, as seen widely deployed during the COVID-19 pandemic.

In this paper, we extend our existing model[1] to incorporate the simulation of arriving travellers having to undergo a period of self-isolation prior to the taking of a post-arrival test. We then look to use our new model to quantify the improvement to detection rates that can be gained by the enforcement of this additional step for isolation periods of varying duration. We again apply this model to simulated outbreaks of COVID-19, influenza, SARS, and Ebola.

## Methods

As this work builds upon our existing model, our core assumptions remain unchanged. The only significant addition being that travellers which managed to board their flight must undergo a fixed period of self-isolation on arrival to their destination, the length of which is pre-defined for each scenario. We consider self-isolation periods of 1 – 14 days.

Similarly, the structure of the model remains largely the same except for the incorporation of the self-isolation period itself. This is implemented by the inclusion of a specification that for each simulated traveller that arrives at their destination and thus enters self-isolation, if their pre-determined incubation period (which is sampled from a distribution particular for each disease) is less than the time from initial infection to the end of their self-isolation, they are assumed to be detected by the test administered at the end of their isolation period. As before, we do not consider disease recovery. Time of exposure (or infection) is sampled from a uniform distribution over a given exposure window (0-3 days, 0-7 days or 0-14 days before departure, representing a short vacation, medium vacation or more longer-term travel). We visualise this new structure below:

**Figure 1:**
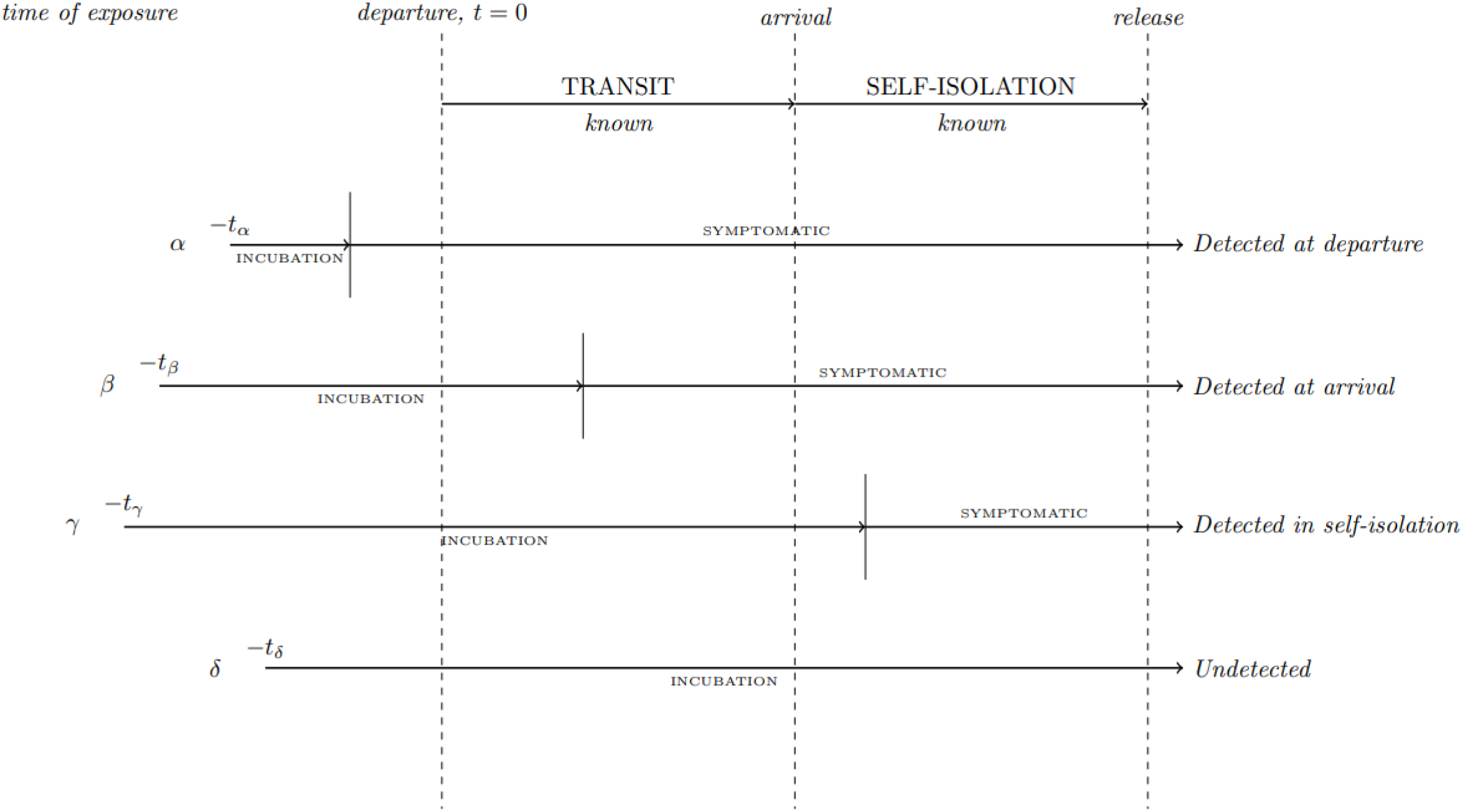
Illustration of the process of evaluating individual simulated travellers in our extended model

The model is once again evaluated across a range of scenarios, each of which is defined by an exposure window, flight type (short, medium or long haul) and the disease being considered. Scenarios are evaluated using a Monte Carlo approach with the detection rate reported being defined as the proportion of infected travellers which managed to travel that were subsequently detected by the end of their self-isolation period (note that we disregard infected travellers detected by pre-departure testing). A more thorough expositions of the model’s underlying process is available in our previous paper[1]. The code use to run this model has been included in the pre-existing Python package (as used in the previous work)which is openly available online[2].

## Results

We table and plot the detection rates as reported by our model for each combination of exposure window and self-isolation period. For conciseness, we have averaged across the results for short, medium, and long-haul flights for each combination. Full results can be found in the supplementary materials.

**Table 1:** Model output for detection rates of considered diseases, exposure window, and enforced self-isolation period. Results averaged across flight type.

|  | Exposure Window (days) | Self-isolation period (days) |  |  |  |  |  |  |  |  |  |  |  |  |  |
| --- | --- | --- | --- | --- | --- | --- | --- | --- | --- | --- | --- | --- | --- | --- | --- |
|  |  | 1 | 2 | 3 | 4 | 5 | 6 | 7 | 8 | 9 | 10 | 11 | 12 | 13 | 14 |
| COVID-19 | 3 | 0.12 | 0.27 | 0.44 | 0.60 | 0.72 | 0.81 | 0.87 | 0.92 | 0.94 | 0.96 | 0.97 | 0.98 | 0.99 | 0.99 |
|  | 7 | 0.22 | 0.39 | 0.55 | 0.68 | 0.78 | 0.85 | 0.90 | 0.93 | 0.96 | 0.97 | 0.98 | 0.99 | 0.99 | 0.99 |
|  | 14 | 0.24 | 0.41 | 0.57 | 0.70 | 0.79 | 0.86 | 0.91 | 0.94 | 0.96 | 0.97 | 0.98 | 0.99 | 0.99 | 0.99 |
| Influenza | 3 | 0.79 | 0.97 | 1.00 | 1.00 | 1.00 | 1.00 | 1.00 | 1.00 | 1.00 | 1.00 | 1.00 | 1.00 | 1.00 | 1.00 |
|  | 7 | 0.79 | 0.97 | 1.00 | 1.00 | 1.00 | 1.00 | 1.00 | 1.00 | 1.00 | 1.00 | 1.00 | 1.00 | 1.00 | 1.00 |
|  | 14 | 0.79 | 0.97 | 1.00 | 1.00 | 1.00 | 1.00 | 1.00 | 1.00 | 1.00 | 1.00 | 1.00 | 1.00 | 1.00 | 1.00 |
| SARS | 3 | 0.12 | 0.27 | 0.44 | 0.61 | 0.75 | 0.87 | 0.93 | 0.97 | 0.99 | 1.00 | 1.00 | 1.00 | 1.00 | 1.00 |
|  | 7 | 0.23 | 0.42 | 0.59 | 0.73 | 0.84 | 0.92 | 0.96 | 0.98 | 0.99 | 1.00 | 1.00 | 1.00 | 1.00 | 1.00 |
|  | 14 | 0.26 | 0.44 | 0.61 | 0.74 | 0.85 | 0.92 | 0.96 | 0.98 | 0.99 | 1.00 | 1.00 | 1.00 | 1.00 | 1.00 |
| Ebola | 3 | 0.00 | 0.01 | 0.02 | 0.04 | 0.08 | 0.14 | 0.21 | 0.30 | 0.39 | 0.48 | 0.57 | 0.66 | 0.73 | 0.79 |
|  | 7 | 0.02 | 0.05 | 0.10 | 0.15 | 0.22 | 0.30 | 0.38 | 0.47 | 0.55 | 0.63 | 0.70 | 0.77 | 0.82 | 0.87 |
|  | 14 | 0.08 | 0.15 | 0.23 | 0.31 | 0.38 | 0.46 | 0.54 | 0.62 | 0.68 | 0.75 | 0.80 | 0.84 | 0.88 | 0.91 |

**Figure 2:**
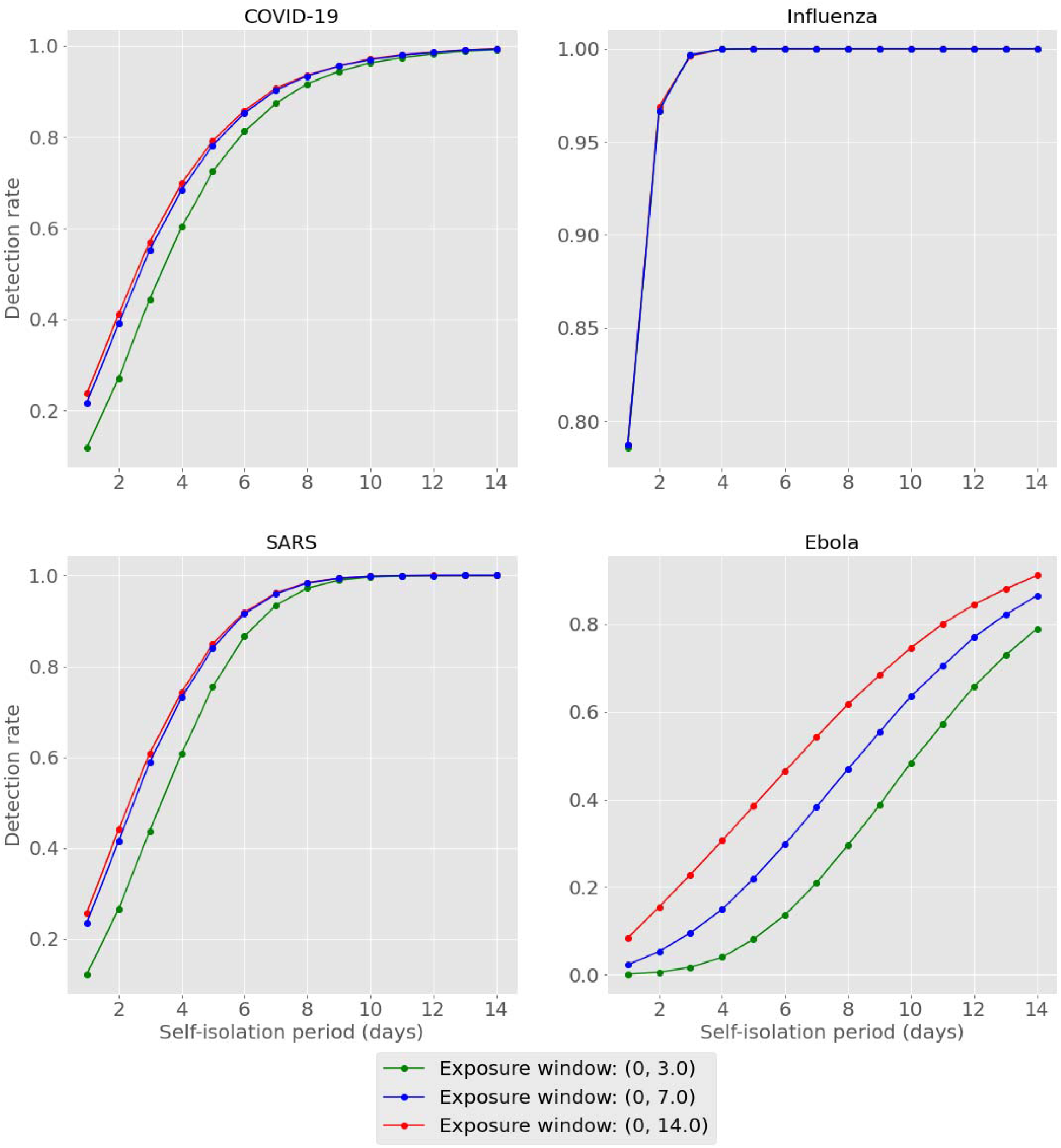
Model output for detection rates of considered diseases, exposure window, and enforced self-isolation period. Results averaged across flight type.

## Discussion

Results show that detection rates increase for all diseases as the length of self-isolation increases. This is in line with the intuition earlier described - allowing infected travellers longer to incubate prior to testing implies a higher chance of cases fully incubating (thus becoming detectable) by the time of testing. Recalling that our previous results suggested detection rates of 9%, 35%, 10% and 3% for COVID-19, influenza, SARS, and Ebola cases in the best case scenario, we see here that these detection rates are at least doubled by the requiring of a self-isolation period of as little as 1 day.

Additionally, we see that enforcing self-isolation yields higher detection rates when controlling for isolation period for diseases with shorter average incubation periods: if we compare influenza to Ebola, we see that by the second day of self-isolation we would expect to be able to detect over 95% of influenza infected travellers versus less than 20% for Ebola in this parameterisation. This suggests that self-isolation periods might be best decided on a disease-by-disease basis, depending on the characteristic of the disease being considered.

Lastly, it should be noted that detection rates seem to become more dependent on the exposure window as the average incubation period increases. Comparing influenza to Ebola again, we see that the detection rates for influenza are almost indistinguishable across exposure windows, while for Ebola we can see that in some cases there is a difference of over 30% in overall detection rate.

In summary, our model indicates that the enforcement of post-arrival isolation period may provide a reasonable method of protecting against international disease importation. However, there is a trade-off between detection rate and the required isolation period, where the latter will have an economic cost in the real world where not all travellers can be expected to be infected. This would be especially true when considering diseases with longer average incubation period (Ebola for example), where the isolation period required to obtain a given detection rate may extend beyond the 14 days considered here.

## Conclusion

In this paper we have described an extension to a Monte Carlo based model we described in previous work. This new version permits the study of the additional protection against case importation that might be gained by requiring travellers to self-isolate on arrival. This model has been applied to simulated outbreaks of COVID-19, Influenza, SARS, and Ebola, where it showed that enforcing a period of self-isolation of any length increases probability of detecting infected travellers. In contrast to our previous work, this extension even suggests that the incorporation of post-arrival isolation might provide considerable protection against international case importation. The methodology used remains flexible so that it may be readily applied to other diseases and scenarios should others wish to so investigate them, thus providing a general tool for international diseases management. The code used herein is an extension of the package used prior and is freely available online[2].

## Data Availability

All data produced are available online at https://github.com/publichealthengland/SIRA

## Notes

### Competing Interest Statement

The authors have declared no competing interest.

### Funding Statement

This study did not receive any funding

## References

[1] D. Bays, E. Bennett, and T. Finnie, “What effect might border screening have on preventing the importation of COVID-19 compared with other infections? A modelling study,” Epidemiol. Infect., vol. 149, p. e238, Nov. 2021, doi: 10.1017/S0950268821002387.

[2] Public Health England, “GitHub - publichealthengland/SIRA,” 2020. [Online]. Available: https://github.com/publichealthengland/SIRA. [Accessed: 09-Jun-2020].

